# Transgender Swimmer in College Athletics

**DOI:** 10.1101/2021.12.28.21268483

**Authors:** Jonathon W. Senefeld, Sandra K. Hunter, Doriane Lambelet Coleman, Michael J. Joyner

## Abstract

There is current scientific and legal controversy about sports competition eligibility regulations for transgender athletes. To better understand and contextualize the effect of androgen-suppression treatment on swimming performance, we compared the gender-related differences in performance of a transgender swimmer who competed in both the male and female NCAA (collegiate) categories to the sex-related differences in performance of world and national class swimmers. These data demonstrate that the gender-related differences in middle distance freestyle performances of a transgender woman are smaller than the observed sex-related differences in performance of top athletes. Our analysis may be useful as a framework for regulators considering participation guidelines which promote fair competition for all athletes, whether cisgender or transgender.

**One-Sentence Summary:** Recent outstanding performances by a transgender woman swimmer are faster than predictions based on historic sex-related differences in swimming performance.

## Introduction

The recent outstanding performances by a transgender woman (male-to-female) swimmer have raised controversy about the role of gender affirming hormone treatments on athletic performance and potential “legacy advantages” of testosterone. Substantial evidence demonstrates that testosterone is strongly associated with sex-based differences in sports performance, more so than any other known factor (*1-3*). The pronounced differences between the sexes in endogenous testosterone levels begin at puberty and correspond to sex-specific divergence in skeletal muscle mass, strength and endurance, cardiac output, lung capacity, hematocrit, and, as a result of this divergence, athletic performance (*1, 2, 4*). This sex-based dichotomy in testosterone is strong evidence supporting endogenous testosterone as a key biological marker to distinguish competitive cohorts in most sports, excluding those sports that are not determined by maximal skeletal muscle or cardiopulmonary performance (for example, chess or electronic sports).

In this context, measuring testosterone levels is the standard practice in many athletic settings. Under the rules of the National Collegiate Athletic Association (NCAA), which governs intercollegiate competition in the US, transgender women who are treated with gender affirming hormones for at least one year may compete as part of women’s sports teams. These rules do not currently distinguish between sports and events, nor do they require that transgender women provide evidence of testosterone levels considered within the normal female range or provide for monitoring for compliance. There is controversy about whether this rule is sufficient to mitigate the so-called legacy effects of hormone— exposure to years of normal male endogenous testosterone concentrations which are, on average, 10 to 20 times greater than normal female testosterone concentrations (*4*). In an effort to determine the extent to which the recent performances by a transgender athlete (with more than 12 consecutive months of gender affirming hormone treatment) might be outliers, we compared the percentage differences in best times of this athlete in the men’s and women’s categories at several freestyle distances to the male to female best time percentage differences of a sample of world and national class swimmers.

The top male and female freestyle swimming performance times at each of six recognized Olympic distances (50, 100, 200, 400, 800 and 1500 m) were obtained, as were corresponding data from NCAA imperial distances of similar durations. Swimming times from long course (50m pool), short course meters (25m pool), and short course yards (25-yard pool) were downloaded from the USA Swimming Database (https://www.usaswimming.org/times/ncaa/ncaa-division-i) for top 25 NCAA times and from the International Swimming Federation (FINA) Database (https://www.fina.org/swimming/rankings) for top 200 World record times. We then paired the male vs. female times (e.g. 1^st^ vs 1^st^, 2^nd^ vs 2^nd^, etc.) within each swimming distance and pool configuration and then calculated a percentage difference for each pair.

After two years of both gender affirming hormone therapy and elite-level swimming training, swimming performance times by a transgender woman were slower by ∼5% across all measured swimming event distances (**Figure 1A**). Despite slower performances, the transgender woman swimmer experienced improvements in performance relative to sex-specific NCAA rankings, including producing the best swimming times in the NCAA for two swimming events (**Table 1**).

**Table 1.** Personal best freestyle swimming performances by a transgender woman (male-to-female transition) in NCAA college athletics in both men’s and women’s competition categories. Personal best performances represent performances from the 2018-2019 NCAA competition season in the men’s category and from the 2021-2022 NCAA competition season in the women’s category. Unranked indicates that the performance was not within the best ∼600 performances of the season. NCAA, National Collegiate Athletic Association.

|  | Swimming Event Distance (yards) |  |  |  |
| --- | --- | --- | --- | --- |
|  | 100 | 200 | 500 | 1650 |
| ***Men's Category*** |  |  |  |  |
| Performance Time (s) | 47.15 | 99.31 | 258.72 | 894.76 |
| '18-'19 NCAA Ranking (No.) | Unranked | 551st | 65th | 32nd |
| ***Women's Category*** |  |  |  |  |
| Performance Time (s) | 49.42 | 101.93 | 274.06 | 959.71 |
| '18-'19 NCAA Ranking (No.)<sup>B</sup> | 132nd | 4th | 4th | 19th |
| '21-'22 NCAA Ranking (No.) | 94<sup>th</sup> | 1<sup>st</sup> | 1<sup>st</sup> | 6<sup>th</sup> |
| ***Categorical Differences<sup>A</sup>*** |  |  |  |  |
| Performance Time (s) | -2.3 | -2.6 | -15.3 | -65.0 |
| Performance Time (%) | -4.6% | -2.6% | -5.6% | -6.8% |
| '18-'19 NCAA Ranking (No.)<sup>C</sup> | -- | +547 | +61 | +13 |

<sup>A</sup>These rankings were interpolated based on season's best performance times in the 2021-2022 NCAA season.
<sup>B</sup>The categorical differences represents the differences in performance of the transgender woman between men's and women's competition categories. See Materials and Methods for further details.
<sup>C</sup>These data were calculated using 2018-2019 NCAA rankings in the men's category and interpolated women's rankings in the 2018-2019 NCAA season. The positive differences indicate that the transgender woman swimmer experienced improvements in performance relative to sex-specific NCAA rankings.

**Fig. 1.**
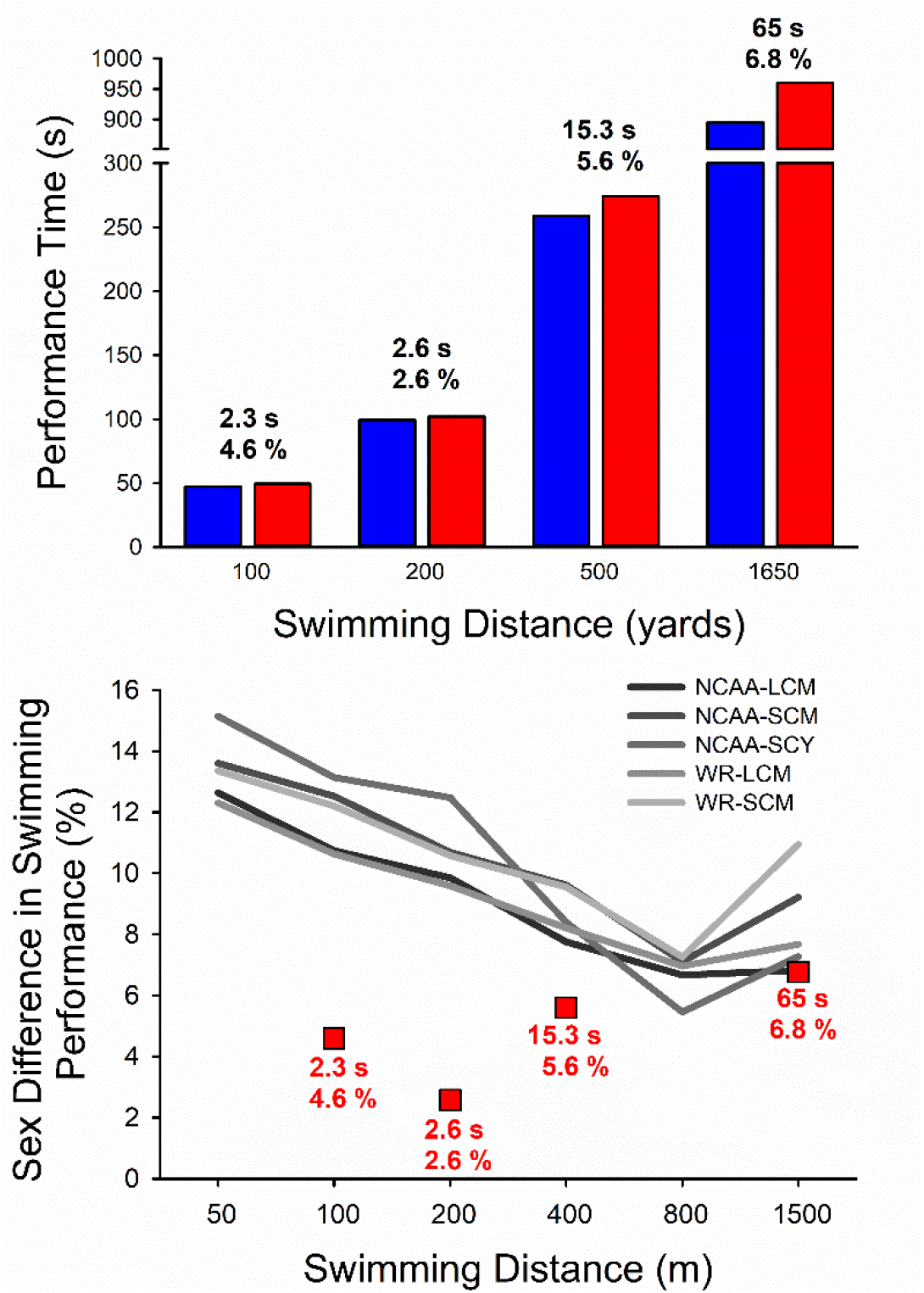
Differences in freestyle swimming performances between men’s and women’s categories for both a transgender woman (male-to-female) swimmer and top swimming performances of all-time in the NCAA and world. **(A)** Bar graph displaying the season’s best performance time of a transgender woman swimmer, including best times from the 2018-2019 NCAA season in the men’s category (blue bars) and from the 2021-2022 NCAA season in the women’s category (red bars). The differences in both performance time and relative performance (%) between the men’s and women’s NCAA categories are delineated in text above each set of comparable bars. **(B)** The grey-scale line charts represent male-vs-female differences in top swimming performances of all-time in the NCAA and world. The differences in performance time of transgender woman swimmer are represented as red squares on metric distances corresponding to imperial NCAA distances. NCAA, National Collegiate Athletic Association; LCM, long course meters; SCM, short course meters; SCY, short course yards; WR, world record.

As a comparator group, we then compared these data to male-vs-female differences in top swimming performances of all-time in the NCAA and world. As expected, based on known physiological differences between the sexes and historical trends, males had faster performances than females in all swimming event distances (*P*<0.001, **Figure 1B**). These differences between swimming performances of males and females were greatest and ∼10-15% for shorter distance events (50 to 200 m) and smaller at ∼7-10% for longer distance events (400 to 1500 m). Of note, these trends were observed in data representing both NCAA-record and world-record performances. Comparing the recent case of the transgender woman swimmer to the historical comparator group of top athletes, we note that her recent performances (compared to her performance times as a man) have smaller differences than comparator data. These data show that for middle distance events (100, 200 and 400m or their imperial equivalents) lasting between about one and five minutes, the decrements in performance of the transgender woman swimmer are less than expected on the basis of a comparison of a large cohort of world and national class performances by female and male swimmers as shown in **Figure 1**.

These analyses were limited by a lack of information on key performance-influencing factors before and after androgen suppression for the transgender woman swimmer (for example, training levels and testosterone concentrations). However, it is possible that the relative improvements in this swimmer’s rankings in the women’s category relative to the men’s category are due to legacy effects of testosterone on a number of physiological factors that can influence athletic performance. In this context, our analysis may be useful as a framework for regulators considering participation guidelines for transgender athletes in the female category. It also raises questions about the duration of the performance enhancing effects of endogenous testosterone after transitioning. Of note, these data present a single case and should not be used to infer definitive effects of gender affirming hormone therapy or legacy effects of testosterone— these topics warrant further investigation.

Accommodations should be made in sports to safeguard fairness for all athletes — whether cisgender or transgender — and will likely vary between sports and events at different levels of competition. While striving for fair and inclusive policies for all athletes, both the magnitude and duration of the profound influence of testosterone on sports performance should be recognized with appropriate consideration.

## Data Availability

All individual data is available in the online databases listed in the text and group data is available in the main text.

## Acknowledgments

None.

## Author contributions

Conceptualization: JWS, SKH, DLC, MJJ

Data Curation: JWS

Formal Analysis: JWS

Methodology: JWS, SKH, DLC, MJJ

Visualization: JWS, SKH, DLC, MJJ

Writing – original draft: JWS, SKH, DLC, MJJ

Writing – review & editing: JWS, SKH, DLC, MJJ

## Supplementary Materials

### Materials and Methods

Sex differences in swimming performance time were calculated for each event distance as: [(male’s performance time) – (female’s performance time)] × (female’s performance time)^-1^ × 100%. All procedures accessed public information and did not require ethical review as determined by the Mayo Clinic Institutional Review Board in accordance with the Code of Federal Regulations, 45 CFR 46.102, and the *Declaration of Helsinki*.

#### Statistical Analysis

Data were reported as means ± SD within the text. Separate mixed-model univariate analyses of variance (ANOVAs) were used to compare the dependent variables (swimming velocity and relative performance (%1^st^ place) of boys and girls, and sex differences in swimming velocity) between three independent variables [age (5-18 years), US ranking (1^st^-100^th^) and event distance (50 m – 1500 m)]. For all other analyses, significance was determined at *p* < 0.05. All analyses were performed with IBM Statistical Package for Social Sciences version 28 statistical package (IBM, Armonk, NY, USA).

